# A New NAEC Level III Epilepsy Center for Adults in Southern San Joaquin Valley of CA – Initial Experience of the Kern Medical Epilepsy Center

**DOI:** 10.1101/2024.05.30.24308251

**Authors:** KT Dao, L Liu, J Ipe, C Poston, N Zalmay, B Ly, C Nguyen, S Braich, C Chacko, J Greenwood, CN Heck, L Ragoonanan, JCT Chen, D Lee, J Russin, B Lee, CY Liu, HP Kunhi-Veedu

## Abstract

Epilepsy care has largely improved across the United States in the past decades, as reflected in Epilepsy Monitoring Units (EMU) bed availability, admissions, neurological procedures, and epileptologists. However, this has not been evident in many underserved areas. The vast majority of National Association of Epilepsy Centers (NAEC) accredited centers are located in major metropolitan areas, and some states lack epilepsy centers entirely. Kern Medical (KM) is the public safety-net hospital in Bakersfield, California (CA) that serves Kern and surrounding counties in the Central Valley. In 2018, an epilepsy center at KM was established with the support of the USC Epilepsy Care Consortium and received NAEC Level III accreditation to serve the over 34,000 patients who suffer from epilepsy in Kern County alone. Here, we review the initial 4-year experience of the KM Epilepsy Center by retrospectively analyzing a prospectively maintained database in light of the general NAEC data from 2012–2019. This experience demonstrates that epilepsy care can be coordinated across complex and competing health systems and socioeconomic barriers separated by large geographic distances through creative, physician-driven strategies of resource sharing and goal alignment across the health care ecosystem. However, considerable challenges remain in providing the requisite care to patients in need, even in highly resourced states like CA. This experience can inform future efforts to integrate epilepsy care across the region and beyond.

## Introduction

Epilepsy is one of the most common neurological disorders worldwide and affects over 3.4 million people in the United States alone. [1] The National Association of Epilepsy Centers (NAEC) has played a critical role in promoting quality epilepsy care by developing standards of care and encouraging their adoption by epilepsy centers across the United States. There has been considerable success, with over 260 specialized epilepsy centers in the USA now having achieved NAEC accreditation. Some states have an apparent abundance of NAEC centers, such as CA (24), NY (24), TX (19), and FL (18). In stark contrast, six states (AK, ME, MT, ND, SD, WY) can count not even one epilepsy center within their borders.[2] Even in states with multiple NAEC centers, the vast majority exist in their metropolitan epicenters. For example, almost all of the NAEC centers in CA exist in the San Francisco Bay Area, Los Angeles Basin, and San Diego, and the majority exist in tertiary care academic medical centers. In the Central Valley ofCalifornia, the population is 6.5 million and rapidly growing, exceeded by only 17 entire states.[3-4] This region is well-recognized to have a large proportion of underinsured patients with many system-level challenges to limit access to complex medical care.[5]

Kern Medical is a public-safety net hospital in Bakersfield, CA that provides critical services to patients in the southern part of the Central Valley, including many who are under-insured. Over 34,000 patients suffer from epilepsy in Kern County alone, and many more utilize the medical resources of Bakersfield. Some patients with adequate resources seek complex epilepsy care in the major metropolitan centers of Los Angeles, San Francisco, and even Santa Barbara, travel many hours to access physicians and resources. A far larger number of under-insured patients rely mainly on state and some federal insurance programs; they have no access to complex multidisciplinary care for epilepsy even if they were willing to travel (Table 1).

**Table 1.**
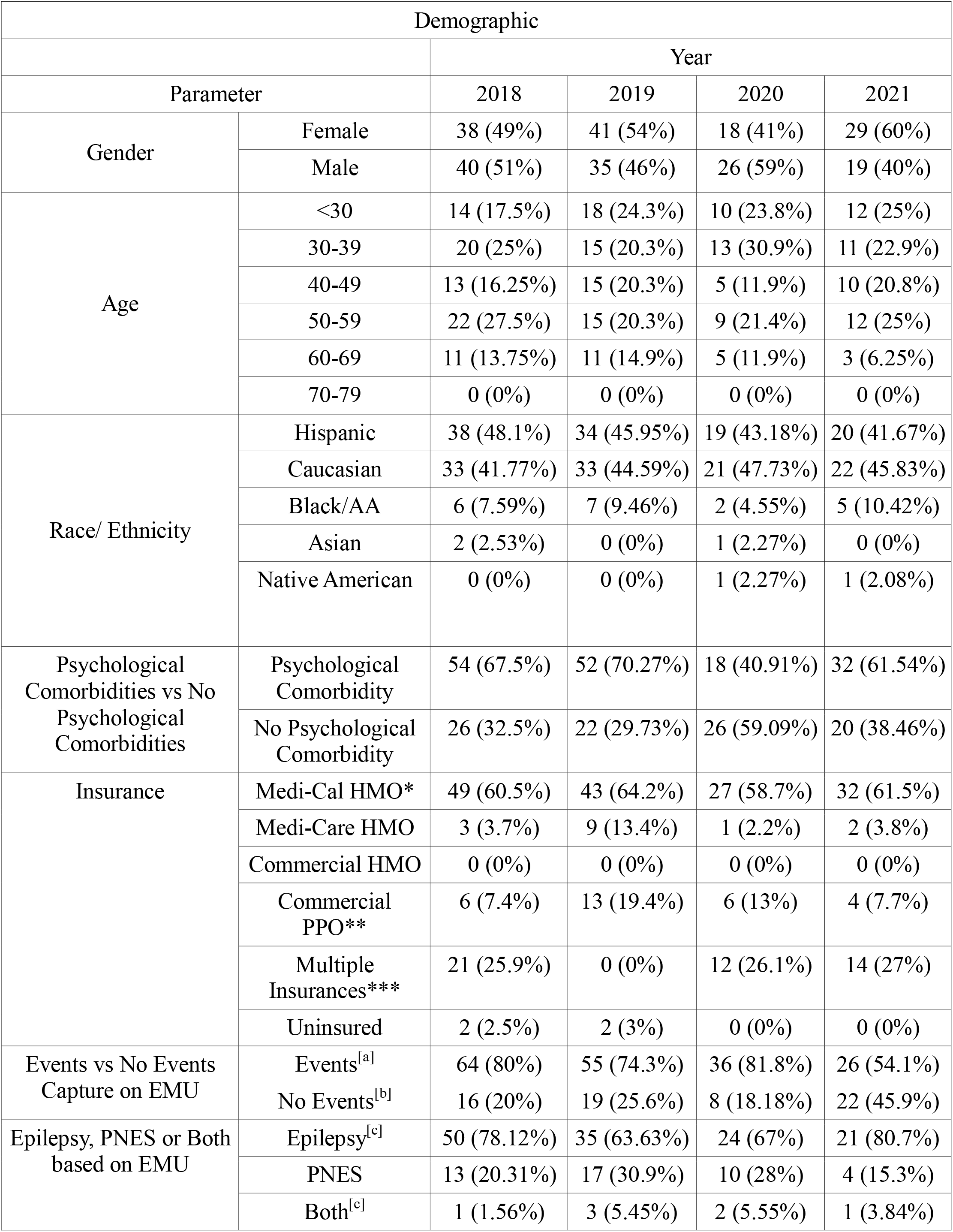
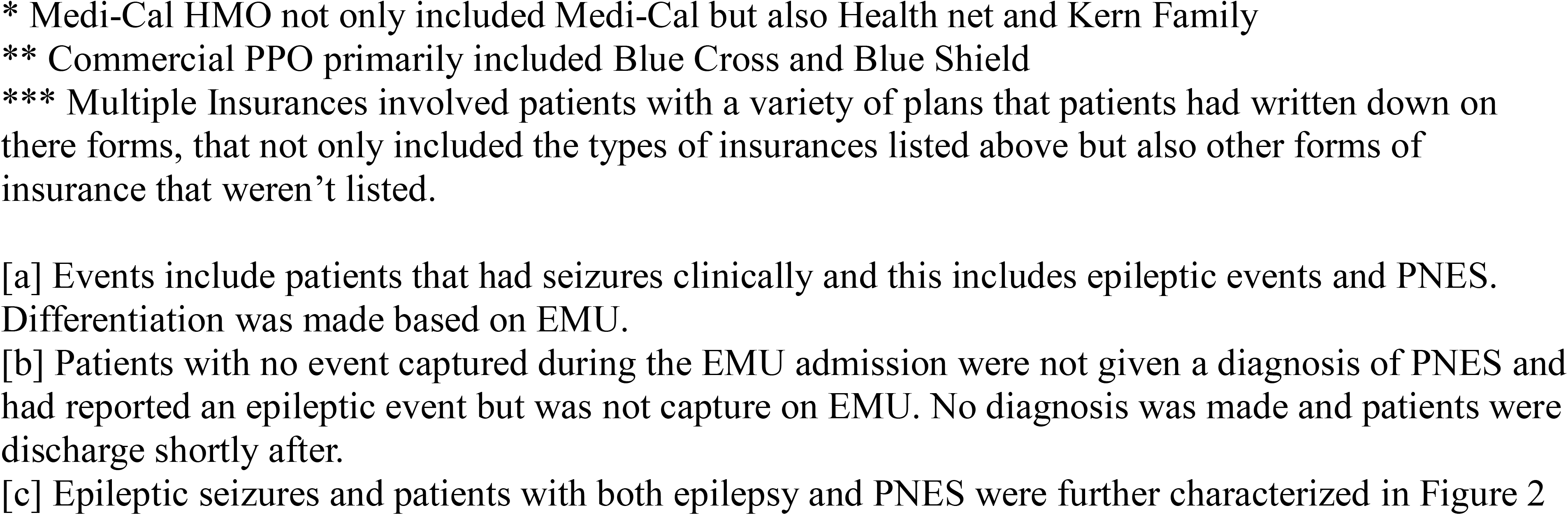
Patient Demographics of Kern Medical Epilepsy Center EMU Admissions from 2018-2021.

| Demographic |  |  |  |  |  |
| --- | --- | --- | --- | --- | --- |
|  |  | Year |  |  |  |
| Parameter |  | 2018 | 2019 | 2020 | 2021 |
| Gender | Female | 38 (49%) | 41 (54%) | 18 (41%) | 29 (60%) |
|  | Male | 40 (51%) | 35 (46%) | 26 (59%) | 19 (40%) |
| Age | <30 | 14 (17.5%) | 18 (24.3%) | 10 (23.8%) | 12 (25%) |
|  | 30-39 | 20 (25%) | 15 (20.3%) | 13 (30.9%) | 11 (22.9%) |
|  | 40-49 | 13 (16.25%) | 15 (20.3%) | 5 (11.9%) | 10 (20.8%) |
|  | 50-59 | 22 (27.5%) | 15 (20.3%) | 9 (21.4%) | 12 (25%) |
|  | 60-69 | 11 (13.75%) | 11 (14.9%) | 5 (11.9%) | 3 (6.25%) |
|  | 70-79 | 0 (0%) | 0 (0%) | 0 (0%) | 0 (0%) |
| Race/ Ethnicity | Hispanic | 38 (48.1%) | 34 (45.95%) | 19 (43.18%) | 20 (41.67%) |
|  | Caucasian | 33 (41.77%) | 33 (44.59%) | 21 (47.73%) | 22 (45.83%) |
|  | Black/AA | 6 (7.59%) | 7 (9.46%) | 2 (4.55%) | 5 (10.42%) |
|  | Asian | 2 (2.53%) | 0 (0%) | 1 (2.27%) | 0 (0%) |
|  | Native American | 0 (0%) | 0 (0%) | 1 (2.27%) | 1 (2.08%) |
| Psychological Comorbidities vs No Psychological Comorbidities | Psychological Comorbidity | 54 (67.5%) | 52 (70.27%) | 18 (40.91%) | 32 (61.54%) |
|  | No Psychological Comorbidity | 26 (32.5%) | 22 (29.73%) | 26 (59.09%) | 20 (38.46%) |
| Insurance | Medi-Cal HMO* | 49 (60.5%) | 43 (64.2%) | 27 (58.7%) | 32 (61.5%) |
|  | Medi-Care HMO | 3 (3.7%) | 9 (13.4%) | 1 (2.2%) | 2 (3.8%) |
|  | Commercial HMO | 0 (0%) | 0 (0%) | 0 (0%) | 0 (0%) |
|  | Commercial PPO** | 6 (7.4%) | 13 (19.4%) | 6 (13%) | 4 (7.7%) |
|  | Multiple Insurances*** | 21 (25.9%) | 0 (0%) | 12 (26.1%) | 14 (27%) |
|  | Uninsured | 2 (2.5%) | 2 (3%) | 0 (0%) | 0 (0%) |
| Events vs No Events Capture on EMU | Events<sup>[a]</sup> | 64 (80%) | 55 (74.3%) | 36 (81.8%) | 26 (54.1%) |
|  | No Events<sup>[b]</sup> | 16 (20%) | 19 (25.6%) | 8 (18.18%) | 22 (45.9%) |
| Epilepsy, PNES or Both based on EMU | Epilepsy<sup>[c]</sup> | 50 (78.12%) | 35 (63.63%) | 24 (67%) | 21 (80.7%) |
|  | PNES | 13 (20.31%) | 17 (30.9%) | 10 (28%) | 4 (15.3%) |
|  | Both<sup>[c]</sup> | 1 (1.56%) | 3 (5.45%) | 2 (5.55%) | 1 (3.84%) |

\* Medi-Cal HMO not only included Medi-Cal but also Health net and Kern Family
\*\* Commercial PPO primarily included Blue Cross and Blue Shield
\*\*\* Multiple Insurances involved patients with a variety of plans that patients had written down on there forms, that not only included the types of insurances listed above but also other forms of insurance that weren't listed.
[a] Events include patients that had seizures clinically and this includes epileptic events and PNES. Differentiation was made based on EMU.
[b] Patients with no event captured during the EMU admission were not given a diagnosis of PNES and had reported an epileptic event but was not capture on EMU. No diagnosis was made and patients were discharge shortly after.
[c] Epileptic seizures and patients with both epilepsy and PNES were further characterized in Figure 2

In 2016, the University of Southern California Epilepsy Care Consortium (USC ECC) initiated a coordinated effort to establish an adult epilepsy center in Bakersfield. A group of experts in epileptology, neurosurgery, neuropsychology, neuroimaging, and support services was assembled by organizing resources in Bakersfield supplemented with those from Los Angeles to open a two-bed epilepsy monitoring unit at KM in January 2018, leading to NAEC accreditation a few months later. [6] Here, we review the initial 4-year experience of the Kern Medical Epilepsy Center (KMEC) over a period that covers the COVID-19 pandemic. We then compare our experience with that of the NAEC national data accumulated from 2012 to 2019 in order to identify ongoing challenges and consider possible solutions to providing complex epilepsy care for all patients in need. [7]

## Patient and Methods

A retrospective analysis was conducted of a prospectively maintained database of the KMEC in Bakersfield, a NAEC Level III accredited center, from 2018–2021. As the first NAEC-accredited adult center with an EMU in the region, it serves an estimated 34,000 epilepsy patients in the Central Valley. From 2018–2021, 246 patients underwent EMU evaluation at KMEC (Table 1). Data collection was conducted primarily through chart reviews and patient interviews. Data collected included: epilepsy diagnosis, medical comorbidities, medical interventions past and present, and medications. The work flow of the KMEC is shown in Figure 1. If patients were deemed to be potential surgical candidates, their case was discussed with a multidisciplinary team consisting of neurosurgeons, epileptologists, radiologists, and neuropsychologists from the USC Epilepsy Care Consortium (Figure 1). Medical management was optimized, and surgical and neuromodulation options were offered if the patients were refractory. In this analysis, we compared each patient’s pre-EMU admission seizure burden with their post-admission seizure burden using the patient’s most recent clinic notes to assess overall benefit. Furthermore, comparisons were made with patients’ pre- and post-seizure diagnoses to determine the impact of EMU admissions on long-term care plans (Figure 1). These values were then used to compare and contrast KMC with the National Association of Epilepsy Centers’ (NAEC) data analysis. [7]

**Figure 1.**
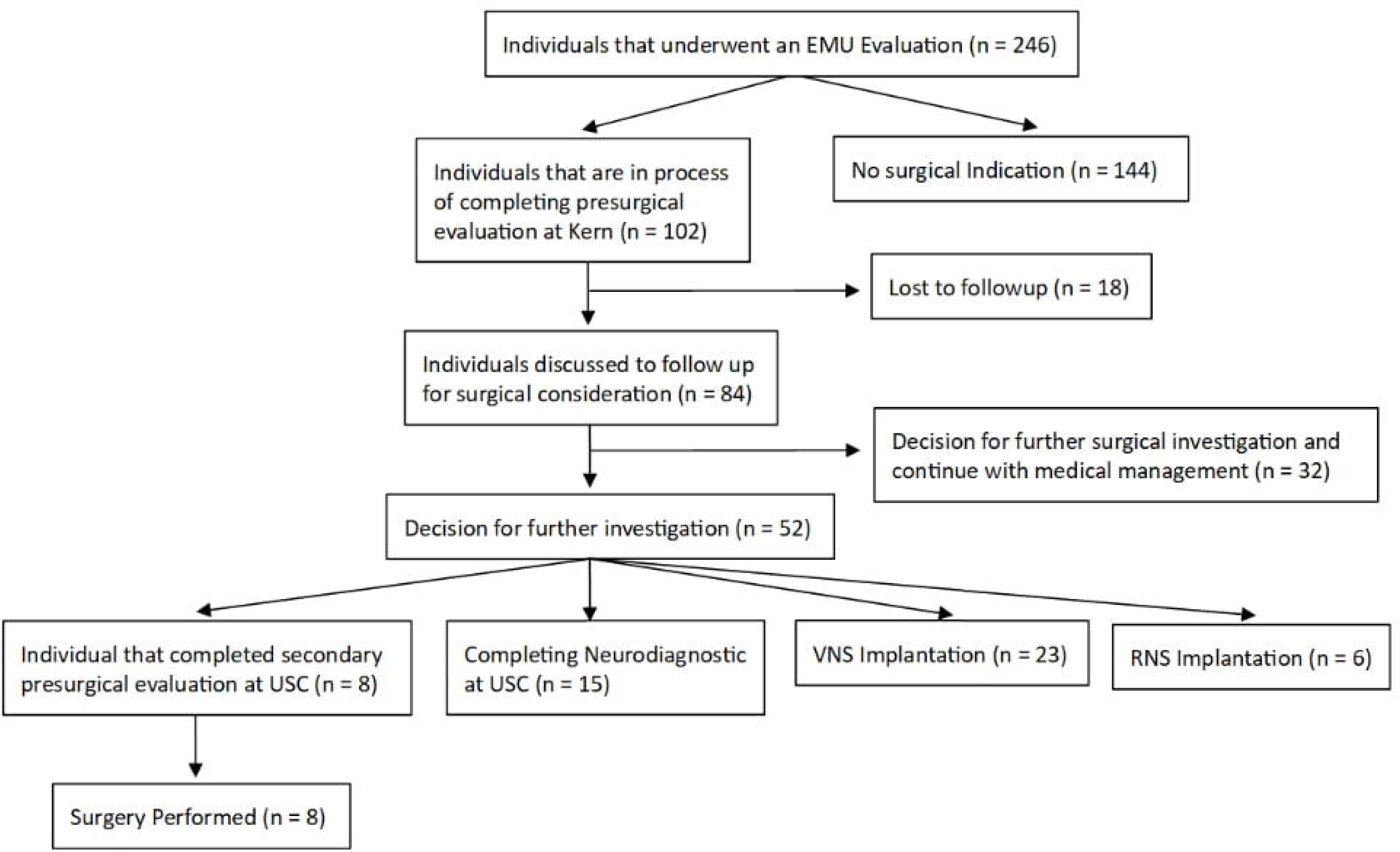
Algorithm of patient who undergo EMU evaluatio11 2018-2021

## KMEC EMU

The KMEC is staffed by 2 epileptologists with a 2-bed EMU (Table 2). Video-EEG monitoring was performed using Nihon Kohden machines. This is a full-array digital EEG systen using a minimum of 19 channels and a maximum of 24 channels for EMU. Disk electrodes were placed on the scalp, and patients were measured according to the 10-20 international system. Frontal and temporal electrodes were used frequently (FT 9, FT 10, and T20 MK). All events were captured through recordings, and any spikes in electrical activity were noted by the seizure detection program. The standard sensitivity is 5-15 uv/mm, with a high filter frequency of 35-70 Hz and a low filter frequency of 0.3-0.6 Hz. Such settings were set at the start of the recordings and adjusted accordingly. Baseline electrode impedance was at or below 10,000 ohms.

**Table 2.** EMU Average at Kern Medical Hospital from years 2018-2021.

| Parameter | Quantity |
| --- | --- |
| Average Number of Epileptologist | 2 Epileptologist |
| Average Number of beds | 2 Beds |
| Average Admissions | 61.5 Admissions |
| Average Length of Stay | 5 nights |
| Average EEG Techs and other staff | 4 other staff members |

All studies were continuously monitored. The events were always detected by recordings but would occasionally be noted by the patient’s caregiver, primary care technician, or nurse. The raw recording data was then transferred from the EEG equipment to a central storage location. An epileptologist, neurologist, and EEG technician would review the entire recording, noting any physical movement and correlating such activity with electrical activity provided by EMU to distinguish epileptic from non-epileptic seizures. An epileptologist would then create the final report.

### Other EEG-Diagnostics

It is well known that epilepsy centers have indirect benefits to medical communities by generally improving all EEG-based services. Thus, we extended our analysis to consider the level of non-EMU EEG-based diagnostic studies done over the same period. These include routine EEG’s, ICU EEG’s, as well as ambulatory video EEG’s. (Table 3)

**Table 3.** Different types EEG Studies performed at Kern Medical Hospital from years 2018-2021.

| Year | Number of Routine EEG | Number of ICU EEG | Number of Ambulatory EEG | Number of EMU Studies |
| --- | --- | --- | --- | --- |
| 2018 | 451 | 153 | 56 | 80 |
| 2019 | 413 | 193 | 69 | 79 |
| 2020 | 376 | 251 | 41 | 45 |
| 2021 | 446 | 335 | 38 | 53 |

## Results

### Patient Characteristics

The characteristics of the patients in the study are shown in Table 1. There was an equal distribution of men and women, with an average of 31.5 females and 30 males. The majority of patients were either of Hispanic descent or Caucasian, and the minority were Black/AA, Asian, and Native American. The average age of patients was noted to be quite diverse, with approximately 54 patients less than 30 years of age, 59 patients between ages 30-39, 43 patients between ages 40-49, 58 patients between ages 50- 59, and 30 patients between ages 60-69. 62.5% of all patients were also noted to have psychological comorbidities such as depression, bipolar disorder, etc., as opposed to 37.5% with no psychological comorbidities.

The majority of the cost of the procedures and EMU studies was covered by patient insurance (Table 1). Kern EMU patients’ insurance information was obtained via interview and electronic medical records, with the majority of the patients using Medi-Cal HMO also known as Medi-Caid HMO with an approximate average of 61.2% of our patient population between 2018 and 2021. Some patients (22%) had documented a variety of different types of insurance. A smaller portion of our patient population also used Medi-Care HMOs, Commercial PPO, or were uninsured. (Table 1). To compare our population to the general population, data regarding insurance information on a state and federal scale was obtained from the KFF and Kaiser Family Foundation websites regarding the health insurance coverage of the total population of the United States [8-9].

### Video EEG Monitor Findings and Diagnostic Outcome

The average length of stay at KMC EMU was 5 nights (Table 2). Throughout 2018, 78 patients underwent EMU monitoring, with 64 events and 14 no events. In 2019, there were 76 patients with 55 events and 21 with no events. In 2020, there were 44 patients with 36 patients with events and 8 without events. Finally, in 2021, there were 48 patients, with 26 having events and 22 having none (Table 1). In patients with no events, a diagnosis was not made. These were patients who reported an event, but none were observed clinically or in an EMU. As such, no diagnosis was made at the time of discharge.

Patients that were diagnosed with a seizure via EMU were then characterized by the type of seizure as well as what could be the underlying cause (Figure 2). Through our findings, we found that 16 patients had generalized seizures and 121 had focal seizures. Patients would then start with medical management initially and be encouraged to undergo additional workups, including imaging such as an MRI, CT, PET, as appropriate, in order to deduce the underlying cause. Patients who were deemed medically intractable would then go through a pre-surgical evaluation, and if they were a good candidate, options for surgery, RNS, or VNS implantation would be discussed (Figure 1).

**Figure 2.**
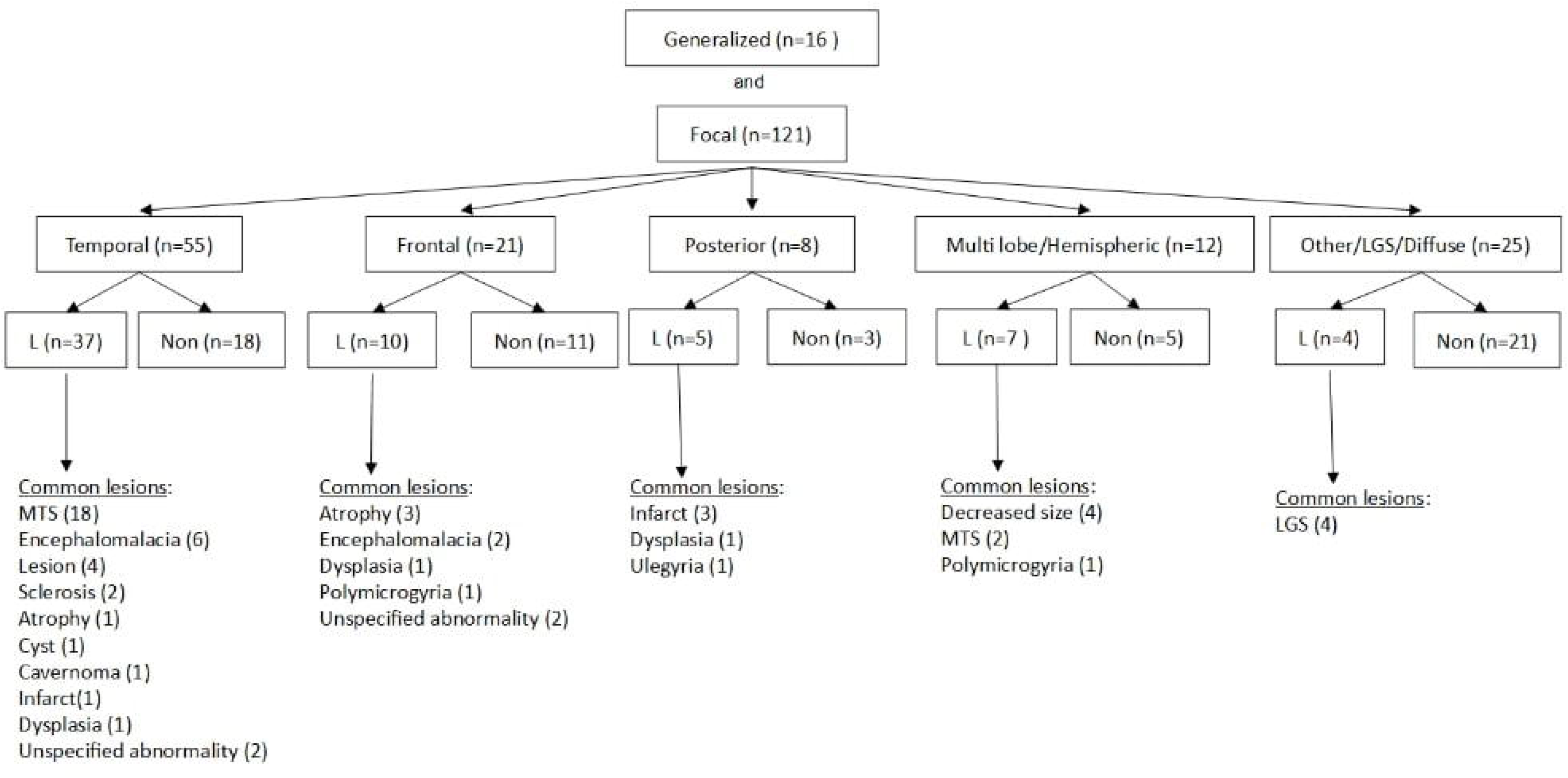
Type of seizure patients were diagnosed with as well as the possible etiology based on MRI^*^.

L = Lesions and Non means no lesion

(LGS = Lennox-Gastaut syndrome)

(MTS= Mesial Temporal Sclerosis Lesion)

^*^This figure includes both epilepsy patients and patients that have epilepsy and psychogenic non epileptic seizures

### Treatment Outcome Surgical vs RNS Implantation vs VNS Implantation

A total of 37 patients underwent surgical intervention over the 4-year period (Tables 4-7), for which there was a noted decrease in seizure frequency. Patients who did not receive surgical intervention received diagnostic clarification and medication management or were lost to follow-up (Figure 1). Patients who were evaluated via our algorithm (Figure 1) had either surgery, RNS implantation, or VNS implantation at a facility that provides a higher level of care (Tables 4-7).

**Table 4.** Kern Medical Epilepsy Center patient procedures 2018-2021.

| Procedure | Number of Patients |
| --- | --- |
| RNS Implantation | 6 |
| VNS Implantation | 23 |
| Surgical Resection | 8 |

Of the 8 patients who underwent resective surgery, 75% achieved significant seizure reduction. Through Neuropace data, it was found that of the 6 patients who had RNS implanted at USC post-EMU stay, 4 experienced >50% improvement in seizure frequency (Table 6). Of the 23 patients who had VNS implantation, only 8 patients experienced seizure freedom or rare and non-disabling seizures while 5 patients had seizure reduction (based upon Engel’s Modified Scoring) (Table 7). [10]

**Table 5.** Seizure burden post-resective surgery after Kern Medical Center EMU evaluation.

| Surgery | Approximate Date of surgery (Month/Year) | Past Seizure Frequency | Current Seizure Frequency | Engel's Criteria |
| --- | --- | --- | --- | --- |
| Left Temporal Lobectomy | 03/2019 | 1-3 times per month | Patient reported a decrease but provided no estimate | IB |
| Right Temporal Lobectomy | 11/2019 | Once a week | None since surgery | IA |
| Right Temporal Lobectomy | 08/2020 | 2-3 times a day | 3-4 times per week | IIIA |
| Right Temporal Lobectomy | 01/2020 | 5 times a month | Once every 6 months | IIB |
| Right Amygdala Cavenoma resection | 11/2018 | 1-3 times per month | Patient reported a decrease and last seizure was 02/2020 | IA |
| Lesion removal of FCD in the right post cortex | 11/2019 | 4-7 times per month | Patient reported a decrease and last seizure was 11/2019 | IA |
| Laser ablation of left anterior temporal lobe | 11/2021 | 2-3 times per week | 2-3 times per month | IIIA |
| Fenestration of left middle cranial fossa arachnoid cyst | 11/2021 | 6-7 generalized tonic clonic seizures in the past year with 2-3 episodes of "deja vu" per day | Occasional Aura's | IB |

**Table 6.** Seizure burden post RNS implantation at USC after Kern Medical Center EMU evaluation.

| Reduction of Seizures | Total Duration of Care |
| --- | --- |
| Patient 1, > 90% Reduction | 8 Months |
| Patient 2, > 60% Reduction | 2 Years |
| Patient 3, > 50% Reduction | 2 Years, 4 Months |
| Patient 4, > 50% Reduction | 9 Months |
| Patient 5, No Change | 3 Months |
| Patient 6, No Change | 9 Months |

**Table 7.** Seizure burden post VNS implantation at USC after Kern Medical Center EMU evaluation.

| Number of Patients (n) | Approximate Seizure reduction |
| --- | --- |
| 11 | Significant seizure reduction of > 50% |
| 1 | Moderate Seizure reduction 5 – 50 % |
| 11 | Little to no noticeable improvement in seizure reduction |

While through the KM EMU and partnership with the USC Epilepsy Care Consortium eliminated the need to repeat most studies when patients were referred to higher levels of care outside of KM, patients a number of barriers have remained. Many treatment options require referring the patients to large academic centers or a level IV epilepsy center. Patients reported they had issues with transportation, payment for treatment, insurance coverage issues, and slow treatment processes.

## Discussion

Similar to that of many parts of the US and the world, epilepsy treatment is highly disjointed. The KMEC was established through the efforts of the large number of physicians that serve patients as part of the USC Epilepsy Care Consortium (USC ECC), which was established at the University of Southern California, Keck School of Medicine to better understand the systems-level issues that lead to a highly disjointed ecosystem for epilepsy care in the southern half of the state of California. Prior to 2018, no NAEC accredited epilepsy center for adults existed in the Central Valley outside the state capitol of Sacramento. The USC ECC now includes8 NAEC accredited epilepsy centers that cover the spectrum of healthcare delivery venues, including academic, community, adult, pediatric, large metropolitan, and small metropolitan hospitals engaged in private sector fee-for-service and public capitated business models. Its members include the USC Keck Comprehensive Epilepsy Center, Children’s Hospital of Los Angeles, Rancho Los Amigos National Rehabilitation Center/USC Comprehensive Epilepsy Center, Children’s Hospital of Orange County Comprehensive Program, Hoag Hospital Epilepsy Center, Cottage Epilepsy Center, Kern Medical Epilepsy Center (KMEC), and Valley Children’s Hospital. This represents one third of all the NAEC accredited centers in CA. Sharing resources, including medical expertise, to allow patients from underserved areas to have access to complex care, remains a central mission of the USC ECC. In addition, the USC ECC fosters the development of independent NAEC accredited centers to provide as much epilepsy care locally as possible. Presently, three additional centers are under development, each strategically located in an underserved geography. Every week, telemedicine conferences are held with participation by all 8 NAEC accredtited member centers, as well as the 3 centers under development. In fact, other epilepsy consortiums, such as the VA Epilepsy Consortium have been creating a network between centers to leverage technology and improve access to care. [11] However, the USC ECC is unique in being a physician-initiated effort to coordinate care across unaffiliated health systems inclusive of shared patient data.

The experience over the initial 4 years of KMEC demonstrates that epilepsy care can be integrated and coordinated across a large geography involving disparate and competing health systems and business models of health care delivery, even in underinsured populations. Compared to the general population, the population treated at Kern Medical has a far greater percentage of Medi-Cal /Medicaid (Table 1). In California, 36.7 % of patients had Medicaid or Medicare as their primary form of insurance, 55.5 % had another type of insurance, whereas 7.8 % were uninsured [8-9]. On a national scale, 34 % of patients had Medicaid or Medicare, 59 % had another type of medical insurance, and 9.2 % were uninsured [8-9]. Many patients that would otherwise not have had access to complex care received critically needed diagnostic evaluation, and many had medication adjustments based on these findings. In addition, a number of patients underwent surgical intervention at KM and other USC ECC partner institutions (mainly USC Keck) with acceptable outcomes. Consistent with the overall NAEC experience, a relatively small number of patients who were admitted to the EMU ultimately underwent surgical intervention. In comparing our experience to national statistics kept by the NAEC from 2012- 2019, we found that more patients have undergone RNS or VNS implantation than epilepsy surgery [7]. The reason why there is a higher number of patients undergoing VNS may be due to cost-effectiveness [12-14], as well as not meeting the criteria for surgery. This is an important consideration given that the majority of our patient population is heavily reliant on government-based insurance (Table 1).

Relative to the national average, the KMEC length of stay is slightly greater. However, the amount of available beds at KMC was half in comparison to the national average with the amount of admissions equating to approximately half or less of all level 3 EMU centers observed (Table 2).[7] Despite KM having a low sample size, the amount of patients requiring some form of surgical intervention was comparable to other Level 3 epilepsy centers nationally (Table 4).[7] Majority of patients who had surgical, RNS, and VNS intervention had some evidence of seizure reduction, with 87.5 % of surgical patients, 66% RNS patients, 52 % VNS patients noting significant decrease in seizure frequency (Table 5-7). However, many patients decided not to undergo further evaluation (n = 144) after discussion with an epileptologist, choosing medical management as first line treatment. (Figure 1) Individuals that had undergone pre-surgical workup but decided not to proceed with surgery also chose medical management (n= 32) after discussions were held between the patient, KMEC, and/or USC ECC. (Figure 1). Most of these decisions were guided by the coordinated workup made possible by the KMEC, highlighting the value of organized epilepsy care far beyond surgical intervention.

Undoubtedly, epilepsy patients admitted through the outpatient pathway have benefited tremendously from the organized approach to EEG-based therapeutics required of comprehensive epilepsy centers. For example, prior to 2018, Kern Medical had no continuous ICU EEG services, a glaring omission given the large number of patients that have neurological disorders, including traumatic brain injury. From 2018 to 2021, the number of ICU EEG’s has more than doubled and far exceeded by 6-fold the number of EMU studies (Table 3).

On another note, the newly established KMEC has also survived the massive disruptions of the COVID pandemic and post-COVID sequelae in the Central Valley. The KMEC continues to function at the same level as the pre-pandemic and has just been re-accredited by the NAEC. In 2022 and 2023, the numbers of ICU EEGs were 434 and 424, respectively. Similarly, the EMU admissions in 2022 and 2023 were 72 and 83.

Regardless of the significant progress that KMC has made over the past few years in treating epilepsy, there remains much more that needs to be done. Despite being among the most advanced economies in the world, epilepsy care in the USA remains highly disjointed, with 6 states not having any NAEC- accredited centers.[2] Until the Kern Level III Epilepsy Center was established, there were no NAEC centers in the entire San Joaquin Valley.[2] This was particularly challenging since over 34,000 epilepsy patients in Kern County alone were deprived of proper epilepsy care. Our experience highlights the ongoing challenges that complex patients face in underserved communities worldwide. Despite an increasing availability of improved medications, invasive diagnostic and therapeutic techniques from laser ablations to neuromodulation, a large number of patients simply have no way to access these treatments. Many patients that require complex surgical treatment evaluated in the KMEC still require surgical treatments found only at the tertiary centers of the USC ECC partners. Clearly, such capabilities must be improved locally. Nevertheless, our experience demonstrates that regional partnerships driven by motivated physicians can at least facilitate a bidirectional patient flow between underserved hospitals and well-established centers, whereby the local centers can provide all the care that is reasonable before referring out, and patients that receive part of their care elsewhere can be easily repatriated to the local providers. The value of shared resources and sharing of patient care data cannot be more valuable to care in epilepsy, particularly when there is considerable geographic distance between the higher levels of care and the patients and families.

The KMEC experience revealed several important points in providing epilepsy care to undeserved areas in the USA. Firstly, we predicted that it would be easier to establish an NAEC center within a public safety-net hospital rather than a private sector hospital, where economic considerations preclude large initial investment and a start-up period. Private hospitals consider referrals outside the system to be undesirable. Within more “mission driven” public sector hospitals, securing the proper resources to establish an epilepsy center, such as EMU beds, EEG techs, dedicated epileptologists and equipment can be justified based on public health considerations, such by raising awareness of the number of patients who suffer from epilepsy in the area. Furthermore, investments can be made with some resources “shared” through partnerships such as exists through the USC ECC. It is of critical importance to design and adhere to a sound business plan, as ongoing subsidies are unsustainable for any health system. At KMEC, the financial health of the program is monitored in an ongoing fashion, and in spite of the adverse health economics of the region, the program is financially sustainable.

Moving forward, many issues remain exceptionally challenging. The financial resources of creating an established center and communicating with complex patients requires many resources, and our experience revealed that, as expected, a majority of patients heavily relied on state government assistance for their surgeries, potentially influencing choice of optional treatments. For example, the majority of the patients who underwent surgical intervention had VNS surgeries. The impact of economics on these decisions is difficult to fully understand and avoid, as they are present in direct as well as indirect ways. This goes to show that access to care is simply the beginning. Another issue that should be considered is that complex cases require a variety of specialists, and patient compliance is greatly affected by language and education, as well as resources to travel to get even a portion of their care that is not available at KMEC.

Despite all the challenges, KMEC was nevertheless established and achieved NAEC accreditation, survived massive disruption due to COVID-19, and it remains sustainable with all the key requirements of an epilepsy center [7,15]. At the minimum, this is a very important demonstration of feasibility. Furthermore, it has served as a useful “living laboratory” to understand the systems-level barriers to integrating epilepsy care in the region, as well as to develop creative solutions moving forward. For example, a much more streamlined bi-directional referral process now exists between Kern Medical and USC Keck specifically for epilepsy, and other members of the USC ECC are now able to bring their resources to bear. Working in close collaboration with USC ECC partner centers was also paramount for our patients.[6] Interestingly, the COVID-19 pandemic left the healthcare delivery infrastructure with far greater capabilities for telemedicine. [16] Communication has never been easier, and virtual meetings allow professionals in various fields to collaborate with increased facility.

## Conclusion

Overall, it is feasible to establish NAEC centers in areas where they have never existed, even in highly underinsured populations. This is greatly facilitated by working in close collaboration with partners in epilepsy care, such as seen with the USC Epilepsy Care Consortium. While feasibility has clearly been demonstrated, there remain considerable challenges that are inherent to the socioeconomics of underserved regions. Finally, careful sustainable growth is critical for ongoing success, with investments made only to gain resources that cannot be easily “borrowed” from collaborating partners where the economics of providing complex neurological and neurosurgical care are more optimal. [17- 18] This can result in a beneficial co-dependency for sustained evolution of the partnerships.

## Study Limitations

We must recognize the limitations of this study, including the retrospective nature and dependency on interviews and telephone contact with patients. Furthermore, the sample size is quite small, with approximately 256 patients across 4 years. Finally, the specific influence of adverse regional economics and patient characteristics cannot be considered in a study such as this. Nevertheless, the study does provide an improved practical understanding of the issues related to integrating the epilepsy care ecosystem.

## Data Availability

All data produced in the present work are contained in the manuscript

## Ethical Approval

Ethical approval to report this case was obtained from the Kern Medical Institutional Review Board; approval ID: 22061

